# Development and optimization of human deuterium MRSI at 3 T in the abdomen: feasibility in renal tumors following oral heavy water administration

**DOI:** 10.1101/2024.12.05.24318155

**Authors:** Mary A McLean, Ines Horvat Menih, Pascal Wodtke, Joshua D Kaggie, Jonathan R Birchall, Rolf F Schulte, Ashley Grimmer, Elizabeth Latimer, Marta Wylot, Maria J Zamora Morales, Alixander S Khan, Huanjun Wang, James Armitage, Thomas J Mitchell, Grant D Stewart, Ferdia A Gallagher

## Abstract

**Purpose:** To establish and optimize abdominal deuterium MRSI in conjunction with orally administered ^2^H-labelled molecules.

**Methods:** A flexible transmit-receive surface coil was used to image naturally abundant deuterium signal in phantoms and healthy volunteers and after orally administered ^2^H_2_O in a patient with a benign renal tumor (oncocytoma).

**Results:** Water and lipid peaks were fitted with high confidence from both unlocalized spectra and from voxels within the liver, kidney, and spleen on spectroscopic imaging. Artifacts were minimal despite the high ^2^H_2_O concentration in the stomach immediately after ingestion, which can be problematic with the use of a volume coil.

**Conclusion:** We have shown the feasibility of abdominal deuterium MRSI at 3 T using a flexible surface coil. Water measurements were obtained in healthy volunteers and images were acquired in a patient with a renal tumor after drinking ^2^H_2_O. The limited depth penetration of the surface coil may have advantages in characterizing early uptake of orally administered agents in abdominal organs despite the high concentrations in the stomach which can pose challenges with other coil combinations.

## Introduction

Deuterium metabolic imaging (DMI) is an emerging method which has shown great promise for non-invasive assessment of tissue metabolism. The technique was first implemented in humans at high field (1), with recent studies showing its feasibility at 3 T (2,3). Although the technique has great potential for imaging metabolism outside of the brain, there have only been a few clinical studies undertaken in the abdomen (1,4,5). Extracranial DMI presents significant hurdles which we have sought to address in this study. We demonstrate the feasibility of abdominal DMI at clinical field strength, which expands the potential applications of the technique.

Deuterium-labelled probes, such as ^2^H-glucose and ^2^H_2_O, are typically administered orally in humans. This poses a physiological problem when using a volume transmit coil for abdominal imaging as acquisition from the organ of interest can be overwhelmed by the signal arising from the stomach, especially early after administration (4). Here we have used a surface transmit-receive coil to minimize the signal from the stomach, while enabling coverage of other abdominal organs. We have compared posterior and lateral coil positions and assessed the kidney following oral ^2^H_2_O administration at 3 T.

On the technical side, the low frequency of deuterium (19.6 MHz at 3 T) poses several challenges. Artifact peaks can arise due to electromagnetic interference (EMI) produced by a variety of sources, such as electronics within the scanning room (motors driving the patient bed, TV screens, CCTV cameras, injection pumps etc.); electronics outside the room but connected through a penetration panel (e.g. gradient amplifiers, narrowband and broadband RF amplifiers); or unrelated RF noise which crosses breaks in the RF cage around the room. Reduction of EMI artifacts is an active area of research relevant to the development of ultra-low field magnets (6). External sources are attenuated via a Faraday cage on conventional clinical MRI systems, and further optimization may be undertaken during system design and installation. However, such optimizations generally focus exclusively on the ^1^H frequency, and significant problems can remain for X-nuclei. In addition, there can be a significant problem with eddy current correction for multinuclear species on some scanners, which we have described previously (7). The magnitude of this problem increases as the difference between the gyromagnetic ratios of ^1^H and the nucleus of interest becomes larger, and therefore for deuterium studies this may be particularly severe. Recently we transitioned to a new MR system for our multinuclear work (GE HealthCare Premier 3 T scanner). Due to all the technical challenges of DMI, it is necessary to characterize these issues and optimize for each new system to reliably implement multinuclear studies.

Here we have shown that these various challenges can be overcome by presenting proof-of-principle images from a patient with a renal tumor following oral administration of ^2^H_2_O. Abdominal DMI at clinical field strength shows promise and could be used for a range of clinical applications in the future.

## Methods

Experiments were performed on a 3 T Premier MR system (GE Healthcare, Waukesha WI). Deuterium image acquisition was performed with a flexible 20 x 30 cm transmit-receive surface coil tuned to the deuterium resonant frequency of ∼ 19.6 MHz at 3 T (Rapid Biomedical, Rimpar, Germany; Fig. 1A). Structural ^1^H-MRI was performed using the inbuilt transmit-receive body coil.

**Figure 1:**
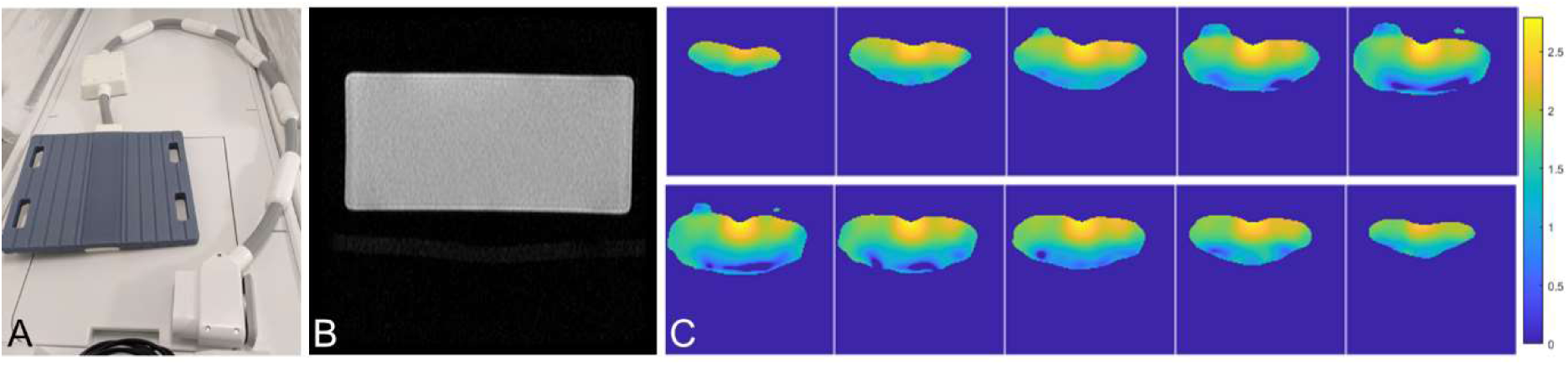
Double-angle B_1_ maps for deuterium. (A) The deuterium surface coil, measuring 20 x 30 cm. (B) A structural ^1^H image of the central slice from a uniform rectangular phantom of silicone oil, dimensions 32 x 22 x 16 cm, FOV 40 cm. (C) A 10 x 10 x 10 matrix was acquired using 3D MRSI with nominal flip angles of 60 and 30 degrees. A 3D B_1_ map was calculated according to the double angle method and interpolated to 128 x 128 x 20. The central 10 slices are shown, covering the 20 cm extent of the coil, which rested on top of the phantom. Intensities represent the ratio of actual to nominal flip angles.

Optimization experiments were performed using two phantoms. Signal strength and uniformity, including mapping of the B_1_^+^ field, were assessed using the naturally abundant deuterium signal within a 32 x 22 x 16 cm rectangular phantom filled with silicone oil (GE Healthcare, Waukesha WI). Spatial resolution and registration with structural images were assessed using the Qalibre System Standard Model 130 phantom (QalibreMD, Boulder, CO, USA), a 20 cm inner diameter sphere of deionized water (H_2_O) which included a plate with a ring of 14 spheres (each with inner diameter 1 cm) containing varying proportions of ^2^H_2_O and ^1^H_2_O (deuterium percentage 0, 10, 20, 30, 40, 50, 60, 65, 70, 75, 80, 85, 90, 95%).

The severity of EMI at the deuterium frequency was assessed by acquiring spectra with the amplitude of the excitation pulses set to zero. Power spectra with 128 averages were plotted with the following sets of acquisition parameters: spectral bandwidth 125,000 Hz, 8192 points, TR 88.7 ms; bandwidth 5000 Hz, 2048 points, TR 419.6 ms.

The severity of spectral distortion due to eddy currents was assessed by acquiring spectra at the minimum TR with no dummy scans (TR 420 ms, spectral width 5000 Hz, 2048 points, 8 transients). The first transient is free of eddy current distortion, while later transients demonstrate effects from the spoiler of the previous TR which immediately precedes the excitation (7). Acquisitions were performed in all 3 orientations in turn, to assess the effect of spoiler application along the X, Y, and Z directions.

Twelve human participants were prospectively included, of which eleven were healthy volunteers recruited to the ethically approved physiological study HeVo-MRI (Yorkshire & The Humber - Leeds East Research Ethics Committee number: 23/YH/0127), and one was a male patient in his 60’s with a left-sided 5.2 cm renal oncocytoma, recruited to the feasibility study IBM-Renal (Ref 8; East of England - Cambridge East REC: 22/EE/0136). Histopathology was determined on surgical specimen post nephrectomy. All participants provided written informed consent. The subjects were positioned supine with the deuterium coil placed either centrally underneath the subjects to target both kidneys (n = 5 healthy volunteers and 1 patient) or wrapped around the right side of the abdomen, secured with velcro straps, to target the liver and right kidney (n = 6).

An axial T_1_-weighted dual gradient-echo fat/water (LAVA-flex) series was acquired in a single breath-hold using the inbuilt ^1^H body coil (flip = 12°, TE = 1.1/2.2 ms, TR = 4.0 ms, FOV = 48 cm, matrix 300 x 200, 64 x 5 mm thick slices, 1 average, 167 kHz full-width receiver bandwidth) for the purpose of anatomical localization. Unlocalized ^2^H spectra were collected from the whole sensitive volume of the coil (TE = 0.7 ms, TR = 600 ms, flip = 90°, 64 averages, total duration = 39 s), followed by Hamming-filtered density-weighted MRSI (10 x 10 x 10 resolution, 1678 transients, FOV = 40 cm, flip = 60°, TE 0.7 ms, TR 250 ms, total duration = 7 min; further details of the trajectory can be found in Figure S1). A matching MRSI dataset with flip = 30° was acquired in 4 subjects for generation of a double-angle method B_1_ map (9). Unlocalized spectra were repeated within the session in 2 subjects, and between sessions approximately 10 days apart in 5 subjects, with a further third session in 3 subjects. The patient was fasted for 2 h prior to imaging and was administered a 5% ^2^H_2_O oral solution (190 g of water for injection + 11.1 g of ^2^H_2_O) immediately before the scan. Healthy subjects did not drink ^2^H_2_O or fast.

MRSI data were zero-filled once in each spatial domain prior to automated peak fitting of water and lipids using the AMARES package (10) within a locally customized version of the OXSA spectroscopy toolbox (11). Voxels were excluded from fitting if the SNR of the maximum peak was below a threshold of 5. The largest peak was assigned to water at 4.7 ppm, and peaks were then fit for water and lipids with initial linewidth estimates of 10 and 20 Hz respectively, with a maximum of 30 Hz. ^2^H_total_ was calculated as the combined signal from ^2^H-water (HDO) and ^2^H-lipids. The ratio of the ^2^H-water to ^2^H_total_ amplitudes were compared in unlocalized spectra and between voxels selected from the liver, kidneys, and spleen. Voxels within the tumor and stomach were additionally compared in the patient. Voxels were excluded from further analysis where the Cramér-Rao Lower Bounds (CRLB) reported for fitting of both peaks was above 10%. Overlays on anatomical images were created using the open-source software ITK-SNAP 3.8.

## Results

### System characterization and optimization

Maps of the transmit B_1_ field in a large uniform phantom showed the expected pattern for a surface coil, with flip angles peaking at the center of the coil and decreasing rapidly with distance (Fig. 1C). In terms of coil penetration, good signal was observed up to a depth of approximately 12 cm, which is similar to the radius of the coil.

The deuterium coil detected many very intense RF spikes in the neighbourhood of the deuterium frequency range due to EMI; however, the 4000 Hz range centered around the resonant frequency of ^2^H_2_O was free of significant artifacts (Fig. 2). The regular pattern of spikes, with the largest ones recurring at intervals of approximately 4000 Hz, suggested an electrical source, but this could not be identified as arising from the lighting or machinery within the scan room.

**Figure 2:**
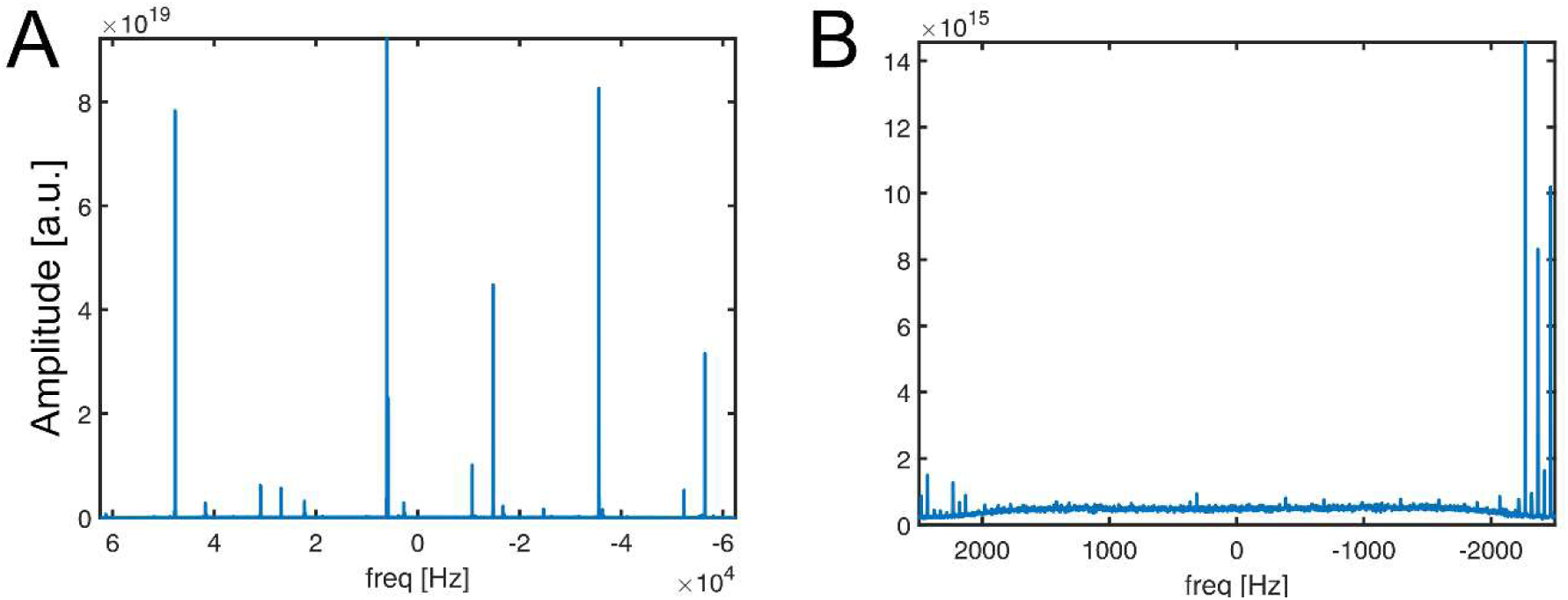
Deuterium spectra acquired with the excitation pulse power set to zero to demonstrate electromagnetic interference. (A) Acquisition over a spectral bandwidth of 125 kHz reveals extensive artifact peaks in the frequency domain. (B) Reducing the bandwidth to 5 kHz, as used for the experiments *in vivo*, demonstrates a lack of significant interference in the center of the spectral range, and a decrease in the maximum peak scale by a factor of approximately 10^4^.

Although the problem with eddy currents was greatly reduced on the Premier system in comparison to a previous GE MR750 system characterized in earlier work (7), it was still present using the initial software version MR29 (Fig. 3). This produced a distortion of the spectral line shapes when acquiring data at the minimum repetition time, and it may also have contributed to a problem we experienced with temporal frequency drift. Over the course of approximately 40 min, the resonant frequency shifted by over 40 Hz, in both the phantoms and *in vivo*. An exponential fit to the curve of frequency shift against time yielded an apparent time constant of 19.4 min. An upgrade to the MR30 software version included our suggested automatic correction for the eddy current problem within the product system software, and following this upgrade, neither line shape distortions nor frequency drift were observed.

**Figure 3:**
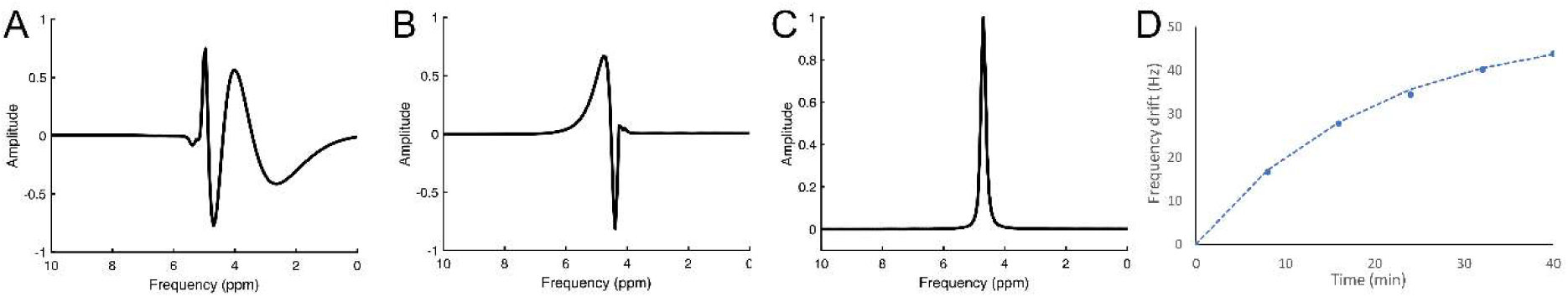
Demonstration of eddy current effects on deuterium data from different scanners and software versions. Distortions in the spectral lineshape as seen on: (A) a GE MR750 device at software level DV26; (B) a GE Premier scanner at software level MR29; (C) the same Premier scanner at software level MR30, following the adoption into the product software of a fix for X-nuclear eddy currents detailed in reference 7; (D) a significant drift in deuterium frequency over time was demonstrated in a static phantom of silicone oil acquired with the MR29 software, which may have been eddy current related as this drift was not present on MR30 software. An exponential fit to the data (dashed line) showed an apparent time constant of 19.4 min.

The Hamming-filtered density-weighted trajectory used for MRSI was shown to have a very narrow point spread function (Fig. S1). Therefore, although the nominal matrix size collected was only 10 x 10 x 10, good spatial resolution could be demonstrated in a phantom (Fig. S2). It was also apparent that a small shift in the left-right direction was needed to align with the structural ^1^H images: this was applied manually in human data.

### Human data

Acceptable fits to unlocalized spectra were obtained in all cases (Table 1 and Fig. 4; Cramér-Rao Lower Bounds (CRLB) for water ranged from 0.9 – 3.2%, mean 1.6%). The CRLB for lipids was always higher than water, especially when the coil was placed laterally rather than posteriorly (mean CRLB 17% lateral vs. 5% posterior, *p* < 0.05). As expected, the linewidths of the lipid peaks were often larger than those of water and were frequently at, or near, the permitted threshold of 30 Hz. The water fraction (^2^H_water_/^2^H_total_) was typically higher when the coil was in the lateral rather than the posterior position, but this was not significantly different (0.86 ± 0.11 vs. 0.75 ± 0.07, *p* = 0.08). The standard deviation of repeated measures of the water fraction within the same session was small (σ = 0.008), but the variation between sessions was higher (average σ = 0.042, range 0.014 - 0.075), which may be secondary to inconsistencies in the coil placement. The lipid fraction (^2^H_lipid_/^2^H_total_) can be derived by subtracting the water fraction from 1 and therefore demonstrates the same standard deviations as the water fraction.

**Figure 4:**
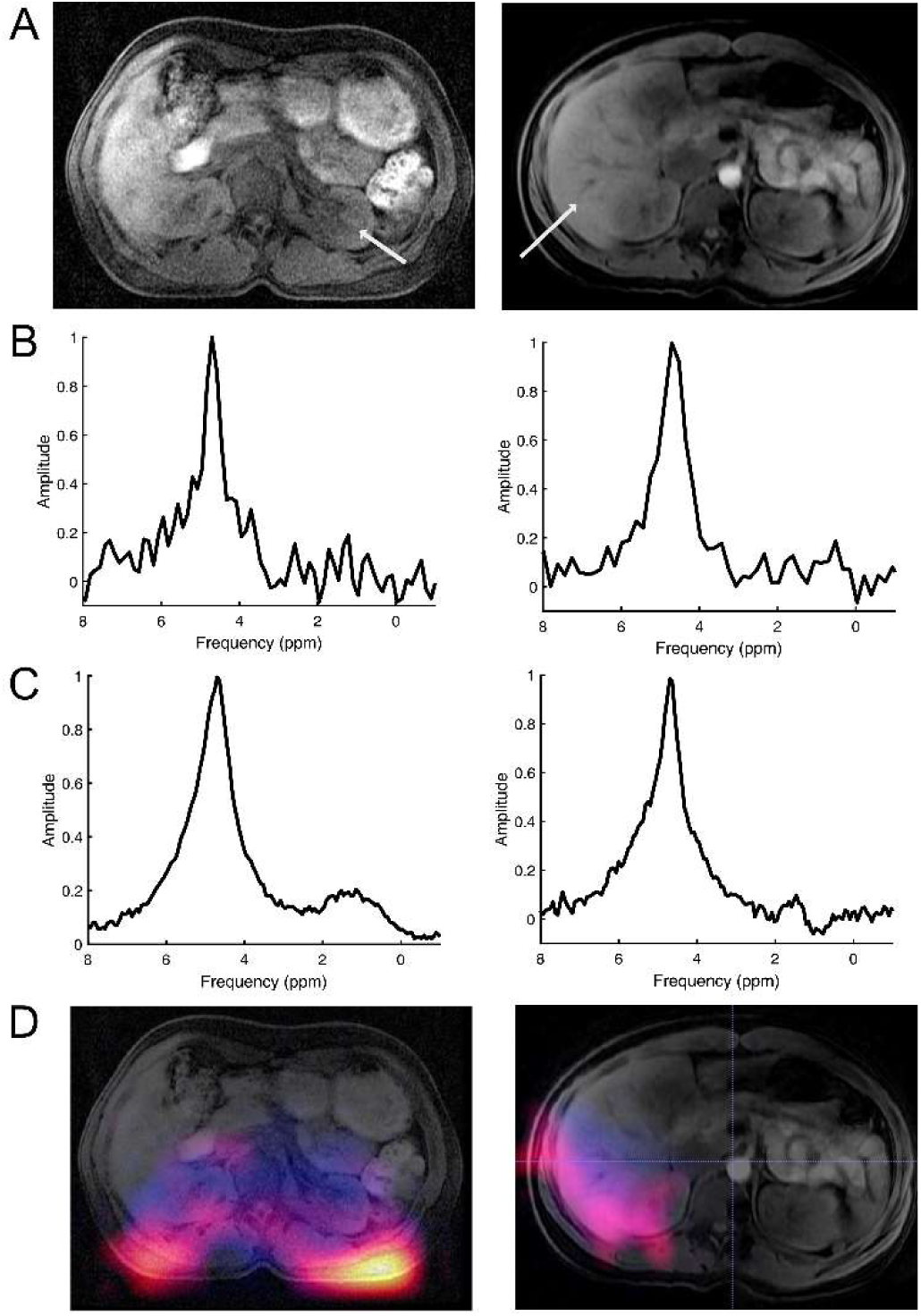
Comparison of data acquired with the deuterium coil placed under the lower back (left column) or wrapped around the right side of the abdomen (right column). (A) T_1_-weighted image showing the location of the extracted MRSI voxel in (B). (C) Spectra from unlocalized MRS. Zerofilling and 1 Hz Gaussian line broadening were applied for display (B and C). (D) Colorscale overlay showing both lipids (“hot” colorscale) and water in blue.

**Table 1:** Data acquired from unlocalized spectroscopy in healthy volunteers.

| Subject number | Coil position | Water $^2\text{H}_{\text{total}}$ | S.D. of repeats | CRB % water | CRB % lipid | Linewidth water (Hz) | Linewidth lipid (Hz) |
| --- | --- | --- | --- | --- | --- | --- | --- |
| 1 | Posterior | 0.80 | N/A | 0.9 | 4.5 | 24 | 30 |
| 2 | Lateral | 0.98 | 0.0077* | 1.4 | 33 | 21 | 2.5 |
| 3 | Posterior | 0.78 | N/A | 1.4 | 4.7 | 29 | 22 |
| 4 | Posterior | 0.80 | 0.0078* | 1.3 | 7.2 | 21 | 30 |
| 5 | Posterior | 0.65 | N/A | 2.0 | 3.1 | 30 | 20 |
| 6 | Posterior | 0.71 | N/A | 2.0 | 5.0 | 30 | 25 |
| 7 | Lateral | 0.85 | 0.058 | 2.0 | 16 | 19 | 30 |
| 8 | Lateral | 0.68 | 0.075 | 3.2 | 7.7 | 26 | 30 |
| 9 | Lateral | 0.92 | 0.022 | 1.5 | 20 | 27 | 30 |
| 10 | Lateral | 0.79 | 0.042 | 1.1 | 5.1 | 26 | 30 |
| 11 | Lateral | 0.92 | 0.014 | 1.2 | 20 | 20 | 30 |
| Mean | | $0.81 \pm 0.11$ | $0.04 \pm 0.02$ | $1.6 \pm 0.6$ | $12 \pm 10$ | $25 \pm 4$ | $25 \pm 8$ |

\*Denotes intra-session repeatability, as opposed to the inter-session repeatability in later subjects.

Acceptable fits were also obtained from spectra extracted from MRSI voxels within the liver, kidneys, and spleen when those were within the sensitive volume of the coil (Table 2). An exception was volunteer 5, where none of the voxels within the kidneys or liver produced SNR above the quality threshold. Ratios of ^2^H_water_ /^2^H_total_ were consistent within and between organs and volunteers. The highest ^2^H_water_ fractions were seen in the liver, but comparisons with other organs did not reach significance (paired t-test comparing liver to the right kidney: *p* = 0.06). Reliable measurements in the left kidney were only achievable with posterior coil placement, as expected given its distance from the coil when wrapped around the right side of the abdomen. Water fractions tended to be higher than in the right kidney, but paired measurements were only obtained in 3 subjects (*p* = 0.15). The standard deviation of repeated measures for the ^2^H_water_ fraction in the liver was 0.06 (range 0.03 - 0.11) and in the kidney was 0.05 (range 0.04 - 0.07).

**Table 2:** Organ-specific ratios of water to total ^2^H signal from MRSI in healthy volunteers.

| Subject number | Coil position | Liver | Right kidney | Left kidney | Spleen | Repeats S.D.: liver | Repeats S.D.: kidney |
| --- | --- | --- | --- | --- | --- | --- | --- |
| 1 | Posterior | 0.84 | 0.83 | 0.85 | 0.80 | - | - |
| 2 | Lateral | 0.86 | 0.76 | - | - | - | - |
| 3 | Posterior | 0.73 | 0.59 | 0.70 | 0.75 | - | - |
| 4 | Posterior | 0.98 | 0.80 | 0.86 | 0.86 | - | - |
| 5 | Posterior | - | - | - | - | - | - |
| 6 | Posterior | 0.75 | - | 0.71 | 0.58 | - | - |
| 7 | Lateral | 0.72 | 0.90 | - | - | 0.105 | 0.036 |
| 8 | Lateral | 0.77 | 0.58 | - | - | 0.032 | 0.046 |
| 9 | Lateral | 0.93 | 0.86 | - | - | 0.075 | 0.068 |
| 10 | Lateral | 0.84 | 0.75 | - | - | 0.035 | 0.056 |
| 11 | Lateral | 0.88 | 0.75 | - | - | 0.048 | 0.040 |
| Mean |  | 0.83 ± 0.09 | 0.76 ± 0.11 | 0.78 ± 0.09 | 0.75 ± 0.12 | 0.06 ± 0.03 | 0.05 ± 0.01 |

After ingestion of ^2^H_2_O, there was a strong water signal detected within the stomach despite the depth of the organ (13-25 cm) which exceeded the usual coil penetration (Fig. 5). However, although this was stronger than the signal derived from other organs at the initial time points, the distance from the coil limited the dynamic range difference and thus prevented severe point spread function artifacts, and this effect disappeared within 30 min. Water signal in the tumor and contralateral kidney remained relatively stable over the time course, while overall unlocalized water signal declined slowly over 40 min (Fig. 6).

**Figure 5:**
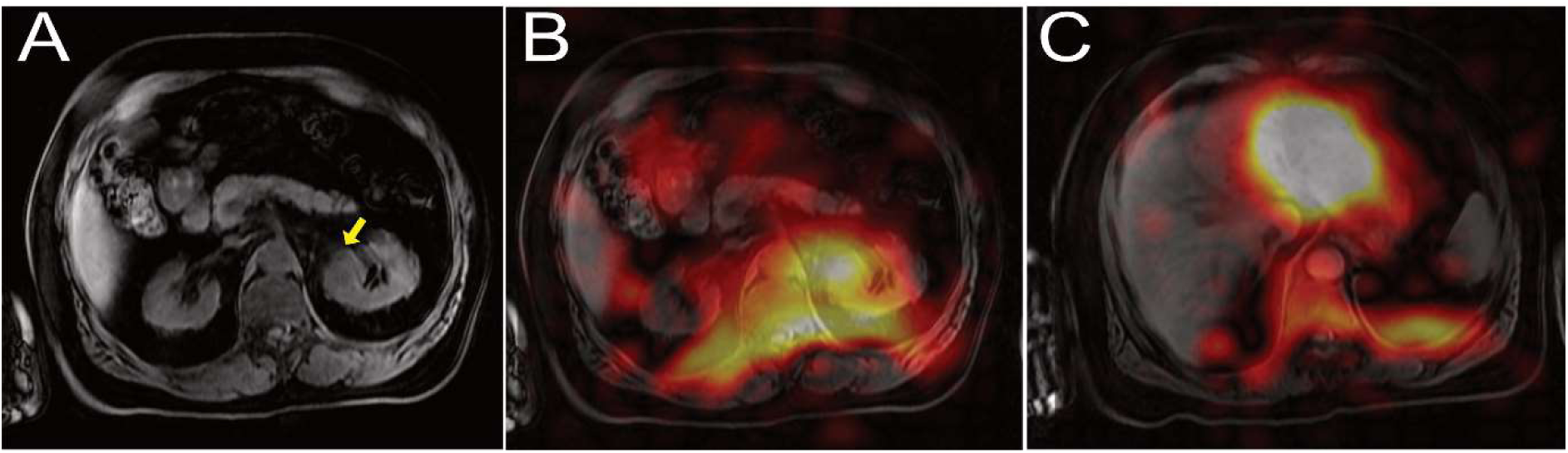
Patient with a renal oncocytoma imaged after drinking ^2^H_2_O. (A) Anatomic ^1^H image at the level of the tumor (arrowed). (B) & (C) Fitted water amplitude, zerofilled to 128 x 128 and overlaid using the ‘hot’ colorscale on the anatomical proton images at the level of the tumor and stomach respectively, 12 min after drinking the labelled water.

**Figure 6:**
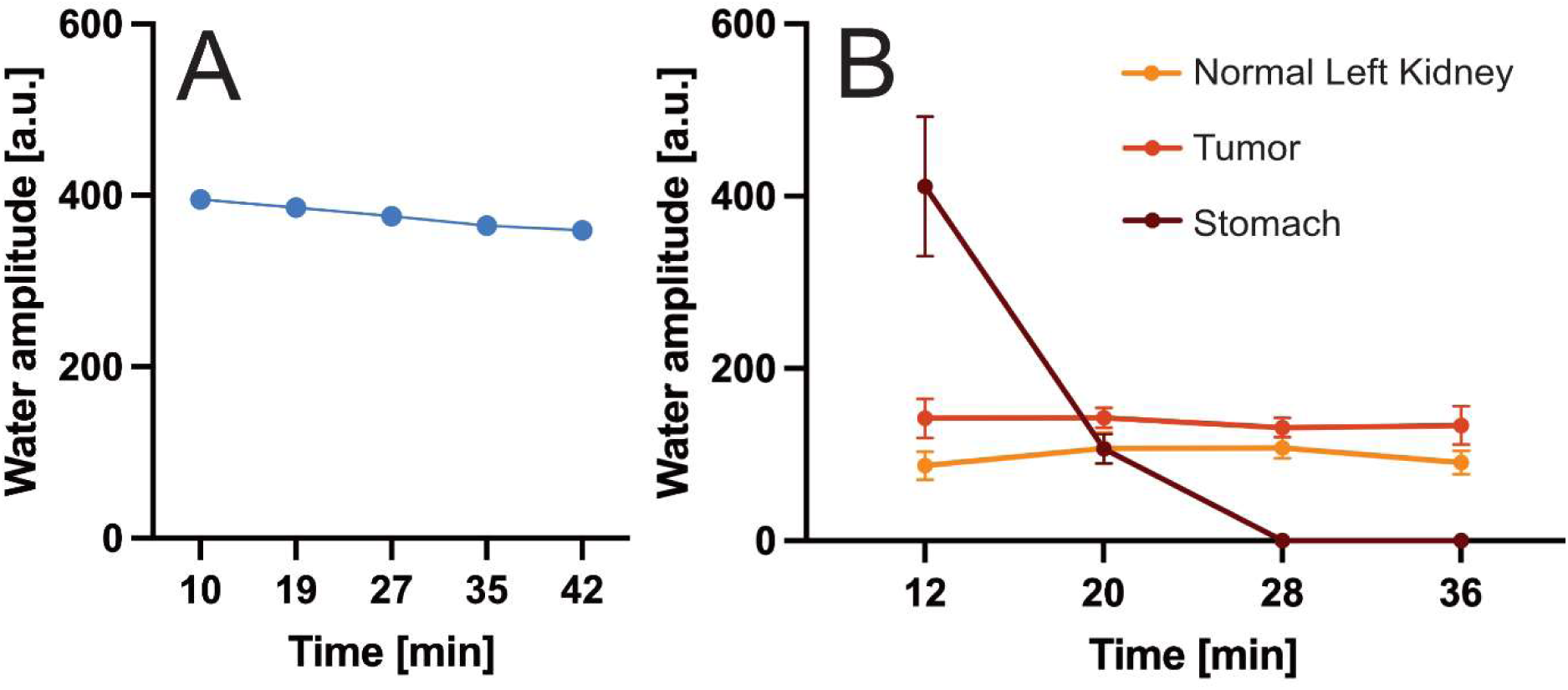
Data acquired from a patient with a renal oncocytoma after drinking ^2^H_2_O. (A) Water peak amplitude in the unlocalized spectra plotted against time from oral ingestion. (B) Water peak amplitude (mean and S.D.) over all voxels within each organ where CRB was <10.

## Discussion

Several technical challenges were overcome to allow implementation of deuterium imaging at 3 T in the human abdomen, both in experimental setup and in characterizing and compensating for data quality issues.

The pattern of electromagnetic interference at the deuterium frequency was characterized and revealed some significant artifact peaks, but fortunately these EMI peaks within 500 Hz of the resonant frequency of ^2^H_2_O were small. Large EMI peaks overlapping the spectral peaks of interest would prevent deuterium imaging and this may be problematic on some systems. Possible solutions include identifying and removing or filtering the sources of the EMI artifacts or ramping the magnet up or down to change the B_0_ field strength and therefore shifting the resonant frequency of deuterium away from the frequencies of the largest artifacts. However, the latter approach could potentially create problems for other nuclei of interest. It should be noted that deuterium imaging techniques which use gradient readouts over a large bandwidth acquisition (e.g. 20 kHz for balanced steady state free precession compared to 2.9 kHz for MRSI (12,13)) are expected to be unworkable on a system which is prone to severe EMI artifacts.

We have previously demonstrated a problem due to an incorrect eddy current correction for multinuclear species on some systems (7): the default correction for the frequency element of eddy current distortion was not correctly scaled for the gyromagnetic ratio, such that the artifact was over-corrected by a factor which increased proportionally to the decrease in the gyromagnetic ratio. Deuterium studies were only possible on that system by applying a compensation to the system calibration files. However, the problem was also proportional to how significant the eddy currents were initially, so it varied considerably between systems. Our GE Premier system produced less severe eddy currents at installation than our previous MR750, but it was still capable of producing a significant distortion of the spectral line shape (Fig. 3). More surprisingly, a drift in deuterium frequency over time was observed, with an apparent time constant of about 20 min. We infer that this also may have been due to eddy currents, because with an upgrade in software version (from MR29 to MR30), which now incorporates the suggested eddy current correction into the default system setup, this frequency drift was eliminated along with the line shape distortions previously observed.

We have previously shown in a DMI study of the human brain at 3 T that the spatial resolution was compromised by using a k-space sampling scheme with a broad point spread function (2). This was deliberately chosen to maximize the SNR, but it may have contributed to the lack of contrast between grey and white matter and the cerebrospinal fluid. The use of such a sampling scheme in the abdomen would be expected to cause severe problems with signal contamination of adjacent voxels, not only from abdominal fat, but also importantly from the signal arising from the orally administered tracer within the stomach. It can be seen in Fig. 5 that although a strong signal from the stomach was seen after ingestion of ^2^H_2_O, it was almost entirely confined to that organ by using a trajectory with a favorable point spread function. Additionally, the signal from the renal tumor can be seen to be distinct from the surrounding kidney, potentially allowing the differences between structures within the abdomen to be quantified.

Traditionally, the use of surface transmit-receive coils has been perceived as undesirable due to their inhomogeneous sensitivity profile; however, some studies have successfully used them for deuterium imaging in the human liver (1,14). Here we demonstrate several advantages of surface coils for abdominal deuterium imaging. The high flip angles near the coil surface, with the ratio of the actual to nominal flip angles close to three, can be exploited to minimize signal from subcutaneous fat. With the nominal flip angle of 60 degrees used in this study, the fat under the skin experiences an actual flip angle close to 180 degrees and is nulled. In the volunteer with the lateral coil placement shown in Fig 3, the unlocalized spectrum shows that virtually no fat signal could be detected.

A more important benefit of using a surface coil is the minimization of signal from the oral tracer within the stomach. According to the Biot-Savart law, for a surface coil of radius R, signal at a depth y is expected to decline on-axis as a function of (y^2^ + R^2^)^3/2^. Since the stomach was at a depth of approximately 20 cm in this subject, the coil receive sensitivity was around 4 times lower than it would have been at a depth of 10 cm approximating to the position of the kidney and tumour. Additionally, the transmit flip angles at that depth would be around 4 times lower for an overall signal reduction of over 13-fold. Even so, the stomach signal in Fig. 6 is more than twice that in the kidney and tumor, showing that if a coil with a homogenous profile were used, this probably would have led to the signal from the stomach swamping the signal from the organs of interest, which demonstrates the potential benefits of the surface coil set up used in this study.

An intravenous injection of the labelled probe, either as a bolus or an infusion instead of oral administration, would eliminate the problem of the hyperintense stomach signal. Several studies of deuterium-labelled probes in tumors and abdominal organs have successfully used this approach in rodents (1,15). Deuterium-labelled glucose for human injection is currently very expensive, although this might be partly balanced by using a smaller dose, and the cost may reduce in time if it is more widely used. A safety consideration with the intravenous use of ^2^H-glucose is the delivery of a large supraphysiological bolus of glucose into the bloodstream and potential side effects from hyperglycaemia, although intravenous glucose can safely be administered as part of a glucose tolerance test to assess for diabetes (16). A large dose of glucose might also potentially alter the steady state metabolism it is attempting measure, and in the brain we have shown that the ratio of non-oxidative to oxidative metabolism was approximately 20 fold higher with an injected tracer (^13^C- pyruvate) compared to an oral agent (^2^H-glucose), albeit using different probes to assess this balance (2). A slow infusion instead of bolus injection may better reflect steady state tissue metabolism (17) but oral administration is ideal for routine clinical use.

Another approach for reducing the impact of the hyperintense signal within the stomach is to wait for several hours between oral administration of the agent and collection of the DMI data. While this may be adequate for determining the distribution of glucose signal in the abdomen, it is unclear whether intersubject variability in glucose absorption from the GI tract might complicate this interpretation. For example, Gursan and colleagues (4) collected data in two subjects with overlapping time courses showing variation between them. In subject 1, the glucose signal appeared to be rising in both liver and kidney 130 min after ingestion, whereas in subject 2, the glucose signal in both organs was already falling at the beginning of the time course, 68 min after intake. These discrepancies could arise from differences in either the rate of glucose absorption or its subsequent metabolism: ^18^F-FDG, the radiolabelled glucose analog widely used in positron emission tomography (PET), is trapped intracellularly unlike deuterated glucose which is rapidly metabolized. The deuterium label could be subsequently transferred into ^2^H-glycogen which is MR-invisible (18), to ^2^H- lactate though glycolysis, or ultimately into ^2^H-water as part of oxidative metabolism (19,20).

Waiting for a prolonged period before measurement is even more undesirable when assessing lactate formation in tumors: as the timescale for the maximal lactate signal following glucose administration is uncertain, observing its change over time is required to accurately assess its production. If a signal were observed at 1.35 ppm in a tumor at a single timepoint some hours after glucose intake, it would be difficult to prove that the signal arose from lactate and not from the overlapping fat signals. We have previously demonstrated using unlocalized whole head spectra in a healthy volunteer that there is a slow steady rise in the peak identified at 1.35 ppm during the hour following glucose ingestion, whereas the fat signal at 0.9 ppm was largely unchanged (2). From this, it is reasonable to infer that an increase in labelling of the lactate pool was the source of the signal elevation. Although in theory it might be possible to observe a decline in such a peak over time some hours after administration due to lactate washout, the time course of that process is likely to be more variable and difficult to interpret.

In healthy volunteers, the coil design allowed us to record naturally abundant deuterium signals from the liver and kidneys. The coil could be placed either posteriorly, for equal sensitivity over both kidneys, or laterally to interrogate the liver and right kidney. A strong unlocalized signal was recorded in all subjects, with a water/fat ratio of around 4:1 on average. Spectral linewidths were approximately 25 Hz, suggesting that the glucose peak at 3.9 ppm will not be fully resolved from water at this field strength. However, our previous work has demonstrated the robustness of spectral fitting using the OXSA spectroscopy toolbox for DMI at 3 T in the brain (2,20). Within-session repeatability of MRS was excellent, with the standard deviation of repeated measures being below 1%. Between sessions, the repeatability was higher, at approximately 5% in unlocalized MRS, and around 7% in the MRSI voxels derived from liver and kidney. These repeated measurements were all made with the coil positioned laterally, where it is more difficult to achieve consistent coil placement. Nevertheless, the repeatability for organ-specific averages was only slightly higher than for unlocalized spectra despite the limited SNR (Fig. 4). This suggests that longitudinal assessments using DMI may be feasible despite the limitation of coil replacement uncertainty.

Although our 7 min MRSI sequence was generally robust, it did not detect sufficient signal from the kidneys and liver for reliable quantification in the subject with the highest BMI (#5) with the coil placed in the posterior position. However, with lateral placement of the coil the liver may have yielded sufficient signal since the layer of subcutaneous fat is thinner over the ribs compared to the lower back and the liver is a more superficial organ compared to the kidneys. Signal could also be improved by increasing the transmit power to achieve higher flip angles deeper in the body, or with longer scan times: collecting 2 averages in 14 minutes should still be clinically acceptable. However, coil penetration remains an obvious limitation of this surface transmit-receive design, particularly in larger subjects.

In a patient with a renal oncocytoma, the deuterium signal was higher in the tumor compared to the surrounding normal-appearing kidney within 12 minutes of oral ingestion of ^2^H_2_O. Importantly, despite the region of interest being located on the left side of the abdomen, it was not overwhelmed by the signal arising from the stomach, which was problematic in previous abdominal DMI studies (4). While we only present a single case of benign oncocytoma in this technical development, future work should also aim to compare signal contrast between benign and malignant renal masses due to the clinical importance of such differentiation and its impact on treatment decisions (8). Recent work in rodents has suggested that perfusion of ^2^H_2_O may be an effective means of producing tumor contrast (21). Changes in ^2^H_2_O uptake related to the effectiveness of cancer therapy have also been seen (22). Although labelling the lactate pool with deuterated glucose would be expected to be a more specific tumor biomarker, the difficulty in distinguishing ^2^H-lactate from ^2^H-lipid might render conventional DMI approaches difficult in the abdomen. Studies with ^2^H_2_O could provide a cheap, rapid, and simple alternative: fitting of the water peak is relatively straightforward and the absorption of water from the gut is much more rapid, reproducible, and less dependent on fasting status than for glucose. Non-calorific liquids are rapidly released from the stomach into the duodenum in an exponential manner, while the presence of nutrients delays gastric emptying depending on the caloric content (23).

## Conclusions

In this study, imaging of naturally abundant deuterium has been demonstrated in the human abdomen at 3 T. Water and lipid peaks were fit repeatably and with high confidence both from unlocalized spectra and voxels extracted from MRSI within the liver, kidneys and spleen.

Repeatability was good within and between sessions, with small variations in the ^2^H_water_/^2^H_total_ ratio observed within and between organs and volunteers. The coil configuration used was shown to avoid excessive artifact from the stomach signal, even within 12 minutes of oral administration of a high dose of ^2^H_2_O. Heavy water may be an interesting probe for tumor imaging and monitoring, and the current work establishes a baseline for future studies using oral deuterated glucose at clinical field strength in cancer and other diseases.

## Data Availability

All data produced in the present study are available upon reasonable request to the authors

## Acknowledgements

This research was supported by Cancer Research UK (EDDPMA-May22/100068, C19212/A27150), the Cancer Research UK Cambridge Centre, the NIHR Cambridge Biomedical Research Centre (BRC- 1215-20014), and the Marmaduke-Sheild Fund. MM acknowledges additional support from the Cambridge Experimental Cancer Medicine Centre, PW from the Gates Cambridge Trust (#OPP1144), JB from the National Cancer Imaging Translational Accelerator (NCITA).

## References

1. De Feyter HM, Behar KL, Corbin ZA, et al. Deuterium metabolic imaging (DMI) for MRI-based 3D mapping of metabolism in vivo. Sci Adv 2018; 4(8):eaat7314.

2. Kaggie JD, Khan AS, Matys T, et al. Deuterium metabolic imaging and hyperpolarized 13C-MRI of the normal human brain at clinical field strength reveals differential cerebral metabolism. Neuroimage 2022;257:119284.

3. Adamson PM, Datta K, Watkins R, Recht LD, Hurd RE, Spielman DM. Deuterium metabolic imaging for 3D mapping of glucose metabolism in humans with central nervous system lesions at 3T. Magn Reson Med 2024; 91: 39–50.

4. Gursan A, Hendriks A, Welting D, de Jong P, Klomp D, Prompers J. Deuterium body array for simultaneous measurement of hepatic and renal glucose metabolism and gastric emptying with dynamic 3D deuterium metabolic imaging at 7T. NMR Biomed 2023; 36(8):e4926.

5. Nam KM, Gursan A, Bhogal AA, et al. Deuterium echo-planar spectroscopic imaging (EPSI) in the human liver at 7T. Magn Reson Med 2023; 90:863–874.

6. Srinivas, SA, Cauley SF, Stockmann JP, et al. External dynamic interference estimation and removal (EDITER) for low field MRI. Magn Reson Med 2022; 87(2): 614–628.

7. McLean MA, Hinks RS, Kaggie J, et al. Characterization and elimination of X-nuclear eddy current artifacts on clinical MR systems. Magn Reson Med 2021;85(5):2370–2376.

8. Horvat Menih I, McLean M, Zamora-Morales MJ, et al. Investigation of the differential biology between benign and malignant renal masses using advanced magnetic resonance imaging techniques (IBM-Renal): a multi-arm, non-randomised feasibility study. BMJ Open (in press) 10.1136/bmjopen-2024-083980, medRxiv 2024.05.03.24306816.

9. Wang J, Qiu M, Yang QX, Smith MB, Constable RT. Measurement and correction of transmitter and receiver induced nonuniformities in vivo. Magn. Reson. Med. 2005; 53:408–417.

10. Vanhamme L, van den Boogaart A, Van Huffel S. Improved method for accurate and efficient quantification of MRS data with use of prior knowledge. J Magn Reson. 1997; 129(1):35–43.

11. Purvis, L.A., Clarke, W.T., Biasiolli, L., Valkovič, L., Robson, M.D., Rodgers, C.T. OXSA: An open-source magnetic resonance spectroscopy analysis toolbox in MATLAB. PloS one 2017; 12: e0185356.

12. Peters DC, Markovic S, Bao Q, et al. Improving deuterium metabolic imaging (DMI) signal-to-noise ratio by spectroscopic multi-echo bSSFP: a pancreatic cancer investigation. Magn Reson Med 2021; 86:2604–2617.

13. Montrazi ET, Sasson K, Agemy L, et al. High-sensitivity deuterium metabolic MRI differentiates acute pancreatitis from pancreatic cancers in murine models. Sci Rep 2023; 13: 19998.

14. Poli S, Emara AF, Lange NF, et al. Interleaved trinuclear MRS for single-session investigation of carbohydrate and lipid metabolism in human liver at 7 T. NMR Biomed 2024; 37:e5123

15. Kreis F, Wright AJ, Hesse F, Fala M, Hu D-E, Brindle KM. Measuring tumor glycolytic flux in vivo by using fast deuterium MRI. Radiology 2020; 294:289–296.

16. Godsland IF, Johnston DG, Alberti K, Oliver O. The importance of intravenous glucose tolerance test glucose stimulus for the evaluation of insulin secretion. Sci Rep 2024; 14: 7451.

17. Hendricks AD, Veltien A, Voogt IJ, Heerschap A, Scheenan TWJ, Prompers JJ. Glucose versus fructose metabolism in the liver measured with deuterium metabolic imaging. Front Physiol 2023; 14:1198578.

18. De Feyter HM, Thomas MA, Behar KL, de Graaf RA. NMR visibility of deuterium-labeled liver glycogen in vivo. Magn Reson Med 2021; 86(1):62–68.

19. Lu M, Zhu XH, Zhang Y, Mateescu G, Chen W. Quantitative assessment of brain glucose metabolic rates using in vivo deuterium magnetic resonance spectroscopy. J Cereb Blood Flow Metab. 2017;37(11):3518–3530.

20. Khan AS, Peterson KA, Vittay OI, et al. Deuterium metabolic imaging of Alzheimer Disease at 3-T magnetic field strength: a pilot case-control study. Radiology 2024; 312(1):e232407.

21. Brender JR, Assmann JC, Farthing DE, et al. In vivo deuterium magnetic resonance imaging of xenografted tumors following systemic administration of deuterated water. Sci Rep 2023; 13: 146999.

22. Asano H, Elhelaly AE, Hyodo F, et al. Deuterium magnetic resonance imaging using deuterated water-induced 2H-tissue labeling allows monitoring cancer treatment at clinical field strength. Clin Cancer Res 2023; 29:5173–5182.

23. Phillips LK, Deane AM, Jones KL, Rayner CK, Horowitz M. Gatric emptying and glycaemia in health and diabetes mellitus. Nat Rev Endocrin 2015;11:112–128.

