## Supplemental Figures for "Development and optimization of human deuterium MRSI at 3 T in the abdomen: feasibility in renal tumors following oral heavy water administration"

Mary A McLean et al.

Submitted to *Magnetic Resonance in Medicine*

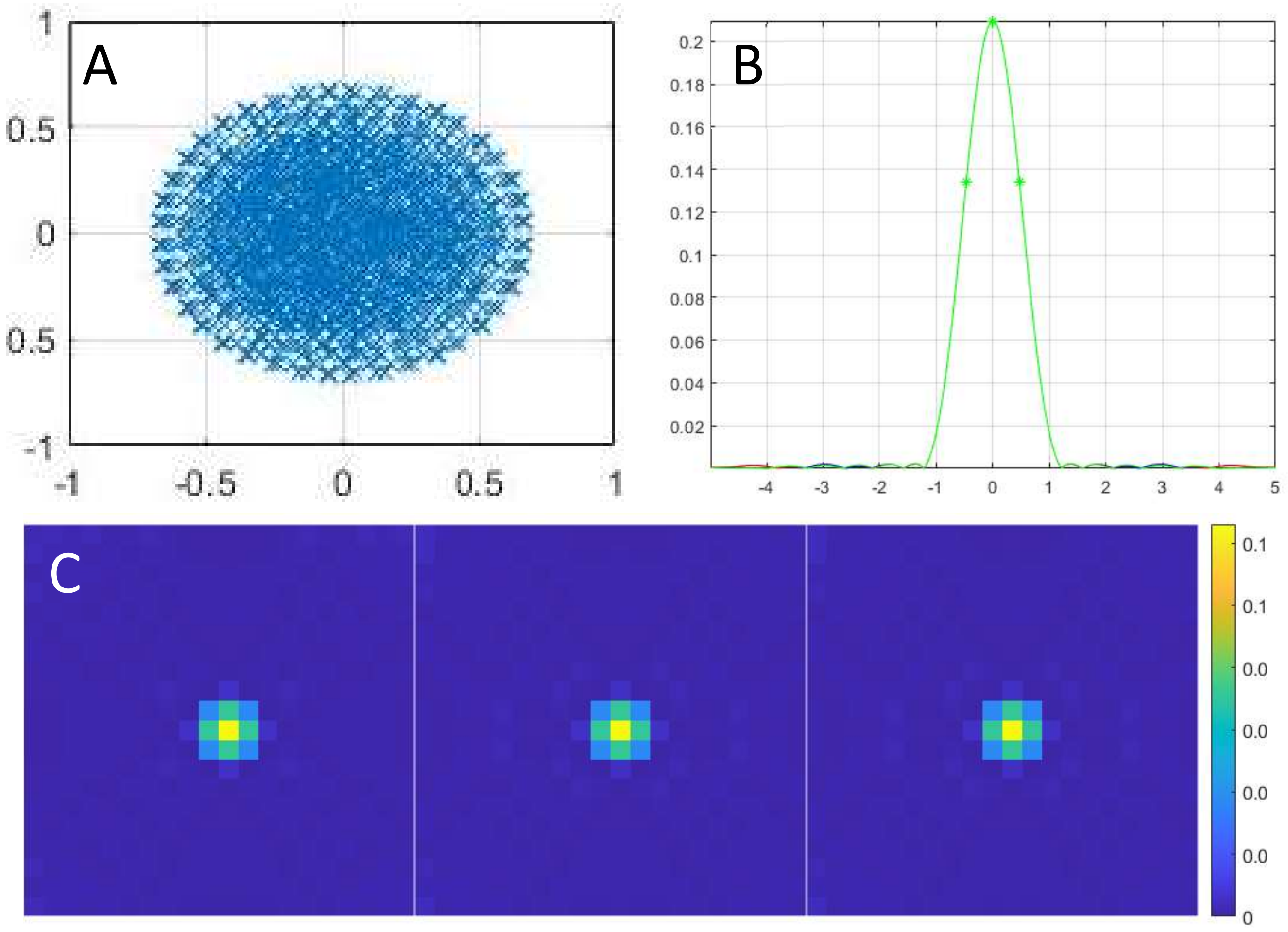


**Figure S1**: Density-weighted MRSI scheme used for acquisition of deuterium metabolic imaging data. (A) Distribution in k-space of the 1678 samples. (B) Calculated point-spread function in each dimension: the asterisks show the full width at 64% of maximum height which is 0.94. (C) Magnitude of the predicted point spread function in the axial, sagittal and coronal directions (shown from left to right).

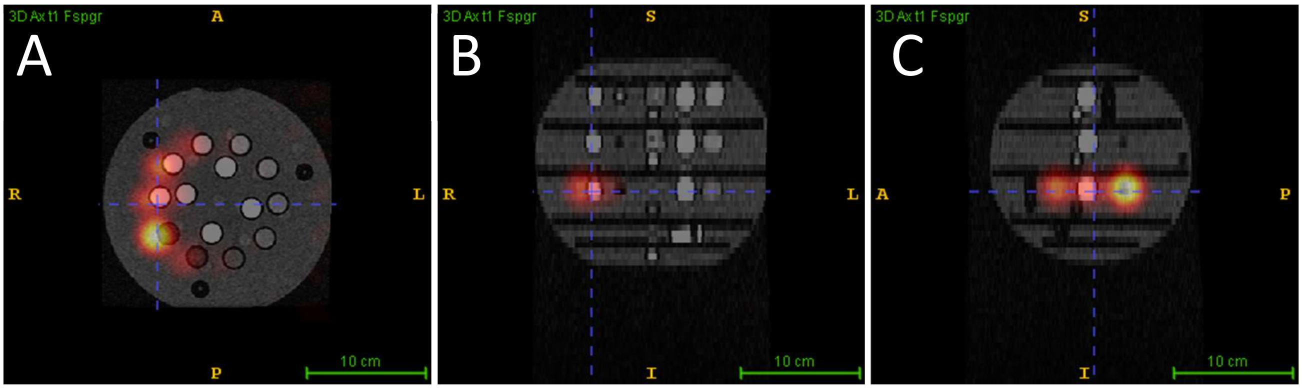


**Figure S2**: Images of peak $^{2}H_2O$ signal intensity derived from MRSI (hot colorscale) overlaid on a 3D gradient echo structural image of the QalibreMD phantom in black and white. Sections are shown in the (A) axial, (B) coronal, and (C) sagittal orientations demonstrating that a shift of the deuterium image toward the right (patient left) is needed to optimally align with the structural image. MRSI was acquired using the same orientation, coil, and trajectory used in the current study.
